# Rapid Point-of-Care Exhaled Breath Analysis for Lung Cancer Diagnosis Using a Micro Gas Chromatography System: A Pilot Study

**DOI:** 10.1101/2024.06.27.24309565

**Authors:** Xingxing Cheng, Yong Feng, Sai Chen, Han Zhang, Ruiping Chen, Bo Xu, Xiao Hu, Wei Wei, Zhenguang Chen, Qian Geng, Junqi Wang

**Author notes:** Correspondence to: Dr. Zhenguang Chen, M.D., Ph.D., Chief Surgeon, Department of Thoracic Surgery, Guizhou Hospital of the First Affiliated Hospital of Sun Yat-sen University, Guiyang, Guigzhou 550003, Department of Thoracic Surgery and Department of Cardiothoracic Surgery of East Division, the First Affiliated Hospital, Sun Yat-sen University, Guangzhou, Guangdong 510080, P. R. China., Qian Geng, M.D, Ph.D., EMBA, ChromX Health Co. Ltd., Greater Bay Area National Center of Nanotechnology Innovation Building, Guangzhou, Guangdong 510555, P. R. China., Junqi Wang, Ph.D., Jingjinji National Center of Technology Innovation, Beijing 100000, and ChromX Health Co. Ltd., Greater Bay Area National Center of Nanotechnology Innovation Building, Guangzhou, Guangdong 510555, P.R. China. These authors contributed equally to this work (Xingxing Cheng, Yong Feng, Zhenguang Chen).

## Abstract

The study investigates the use of volatile organic compounds (VOCs) in exhaled breath as a non-invasive diagnostic tool for lung cancer (LC). Employing a novel micro gas chromatography-micro photoionisation detector (μGC-μPID) system, we aimed to identify and validate VOCs that could differentiate between LC patients and those with benign pulmonary diseases. The cross-sectional study included 106 participants, categorized into 85 LC patients and 21 benign controls, based on computed tomography and histological assessments. Participants provided breath samples following a standardized protocol, and the μGC-μPID system, known for its rapid point-of-care capabilities and low detection limits, was utilized for rapid and sensitive online VOC analysis. Through a meticulous process of data analysis, including principal component analysis, single-factor hypothesis testing, orthogonal partial least squares discriminant analysis and various tests of machine learning algorithms, including random forest, k-nearest neighbor, logistic regression, XGBoost, and support vector machine, we finally identified six potential VOC biomarkers, with diagnostic models incorporating these markers achieving high sensitivity (0.95-1.00) and specificity (0.84-0.88), and areas under the receiver operating characteristic curve ranging from 0.79 to 0.91. Moreover, these models were also extended favourably to the recurrence and metastasis of pulmonary cancer and oesophageal cancer. The study demonstrates the potential of μGC-μPID as a point-of-care tool for LC differential diagnosis, highlighting the need for further validation in larger, multi-centric cohorts to refine the VOC biomarker panel and establish a robust diagnostic framework for clinical application.

## 1. Introduction

Lung cancer (LC), arising from the bronchial mucosa or glandular elements, has emerged as the preeminent etiology of cancer-induced mortality on a global scale. The year 2020 witnessed a staggering 2.22 million incidences and 1.80 million fatalities attributed to LC, constituting approximately 20% of the total cancer-related deaths [1]. Notably, the prognosis for advanced-stage LC patients, is dishearteningly poor, with survival rates only ranging between 10 to 20% [2]. This stark reality underscores the imperative for the early identification of the disease and the stratification of individuals at elevated risk, thereby enabling the deployment of targeted therapeutic interventions and management protocols. Despite advancements in diagnostic techniques, the prevailing methods, predominantly rooted in imaging technologies, present formidable challenges including suboptimal sensitivity, challenges in discriminating between benign and malignant nodules, an inability to detect diminutive lesions, and the financial burden and ionising radiation hazards [3]. Moreover, histopathological investigations are invasive in nature and thus ill-suited for broad-based early screening of populations at risk. Consequently, there exists an exigent demand for the innovation of diagnostic strategies that are non-invasive, economically viable, and characterized by a high degree of precision, with the ultimate aim of augmenting early detection and markedly enhancing therapeutic outcomes [4].

Over the past several decades, the examination of volatile organic compounds (VOCs) present in exhaled breath has attracted considerable attention as a potential source of biomarkers for pulmonary afflictions including LC [5]. Exhaled VOCs, either endogenous or exogenous, are gaseous organic molecules with a high vapour pressure at environmental temperature and a boiling point typically ranging from 50 to 250 °C. The appeal of exhaled VOCs as diagnostic indicators lies in their non-invasive acquisition, simplicity of collection, and capacity to mirror changes in pathogen proliferation or the host’s immune response [6].

Recent studies have illuminated the potential of breath VOCs as biomarkers for the early identification and differential diagnosis of LC [7–27]. Gordon et al. [28] pioneered the use of gas chromatography-mass spectrometry (GC-MS) to identify alkenes in the breath of LC patients. Subsequently, Phillips et al. [29] demonstrated that a panel of 22 exhaled VOCs could effectively distinguish between individuals with and without LC before their further investigation [30, 31]. A Polish research group conducted a series of studies, meticulously analyzing GC-MS profiles to identify 19 to 32 VOCs at parts per billion (ppb) levels, specific to various LC subtypes, including small cell LC (SCLC) and non-SCLC [32, 33]. Peled et al.’s comparative analysis using GC-MS revealed significant differences in 1-octene concentrations between malignant and benign tumor patients. Employing an electronic nose, they achieved a sensitivity of 86% and specificity of 96% in group differentiation [27]. Broza et al. [18] developed a diagnostic model with a sensitivity of 100% and specificity of 80% based on a sample of 10 patients with benign tumors and 24 with LC. Fu et al. [26] observed increased levels of specific VOCs in the breath of LC patients compared to healthy controls (HCs) and those with benign tumors. Corradi et al. [34] developed a classification model with a sensitivity of 60.6% and specificity of 67.2% for distinguishing between patients with LC and those with benign tumors. These findings significantly advance the field of breathomics and underscore the potential of breath VOC biomarkers for non-invasive LC diagnostics and highlight the imperative for further optimization and validation of these VOCs to refine the sensitivity and specificity of breath analysis in the context of LC detection and management.

Notably, most research to date has employed GC-MS and electronic nose (eNose) for VOC biomarker identification or breath-print pattern recognition [35]. However, GC-MS requires complex and time-consuming preconcentration procedures rendering it unsuitable for clinical practice while eNose is typically designed for targeted VOC detection with lower sensitivity and no capacity for identifying potential biomarkers [36]. In this study, we used a novel portable online micro gas chromatography-micro photoionisation detector (μGC-μPID) system to detect breath VOCs for LC distinguishing from benign diseases. μGC-μPID is a rapid point-of-care (POC) breath VOC analyser [37] previously applied successfully in breath analysis of COVID-19 [38], asthma [39], acute respiratory distress syndrome [40, 41] and colorectal cancer (CRC) [42], previously. We demonstrate here for the first time the potential application of μGC-μPID in practical LC differentiating diagnosis in computed tomography (CT) abnormal patients.

## 2. Methods

### 2.1 Study design and participants

This cross-sectional study, conducted from November 2021 to January 2022 at the East Division of the First Affiliated Hospital of Sun Yat-sen University in Guangzhou, China, received approval from the Ethics Committee of the First Affiliated Hospital of Sun Yat-sen University (No. 2022-016). Prior to the initiation of clinical trials, all subjects provided signed informed consent. Subjects, categorised into LC patients and benign controls (BCs), were breath-sampled. This categorisation was based on CT and histological examinations. All enrolled patients had not previously received any malignancy-related treatment and presented histological lesion results. The age range of all participants was 18-80 years. Exclusion criteria included unwilling or unable to sign informed consent in person, unqualified breath sample, relapsed diseases with incomplete treatment history, suffering from other malignant tumors, with severe bronchial asthma and confirmed tuberculosis or with severe liver damage and kidney diseases. Recording of demographic and clinical information were collected.

### 2.2 Breath collection

Breath sampling operators underwent professional training to ensure standardization of procedures. All subjects were asked to fast for at least 2 hours, rinse their mouths with purified water, and abstain from vigorous exercise, alcohol consumption, and smoking before breath collection. Exhaled air in 3 min was directly pumped into the gas inlet of μGC-μPID for VOC analysis (about 600 mL were collected) while all the rest breath air was collected into a pre-processed sorbent tube simultaneously. All breath samples were collected in the same room during this study. Each sorbent tube was sealed at both ends with brass caps containing polytetrafluoroethylene ferrules. Prior to use, the sorbent tubes were aged at 320°C for four hours while purged with nitrogen of 99.99% purity.

### 2.3 μGC-μPID analysis

The μGC-μPID system (ChromX Health Ltd., China) comprises three distinct, silicon-based microfabricated chips: a multi-adsorbent packed micropreconcentrator-injector (μPCI) for VOC capture, preconcentration, and injection; a 10 m long microcolumn integrated with thin-metal heaters and temperature sensors for temperature-programmed separations; and a microfabricated photoionization detector. In a full analysis cycle, the breath sample was directly drawn through a Nafion tube to remove moisture, then through the μPCI at a fixed flow rate using a mini pump. The captured VOCs were then injected into a μcolumn by a rapid thermal desorption (∼300 ms). In the column, the VOC mixture was separated under conditions of a 1 mL/min carrier gas flow rate and a temperature programme with a ramp rate of 10 °C/min from 25 °C to 180 °C.

Additionally, the sorbent tube is connected to a μGC-mass spectrometry device (MSD) for further VOC identification. This complete platform includes a high-throughput automatic injector, a homemade thermal desorber, a μGC-μPID, and an MSD (Agilent 5977B). The tube undergoes thermal desorption under standard settings: a flow path at 180LJ, and a pre-purge at 100 mL/min for 2 min to remove water moisture. The sample tube is desorbed at 300LJ for 10 min, with the flow rates set at 60mL/min. Mass spectra were obtained using Qualitative Analysis 10.0 (MassHunter) software and cross-referenced with the NIST 2017, Version 2.3 mass spectrum library.

### 2.4 Data analysis

Demographic characteristics and VOC peaks across different groups were compared using the independent *t*-test for data with normal distribution and the rank sum test for non-parametric data in univariate analyses. Statistical significance was set at *p* < 0.05, and the Benjamini-Hochberg procedure was applied to all *p*-values to calculate the false discovery rate (FDR) value.

Raw data from μGC-μPID were initially processed to eliminate noise and offset errors generated during data collection procedures, including baseline correction, noise cancellation, and time alignment. Subsequently, a peak detection algorithm was applied to the aligned data to identify each peak and its corresponding metabolite, facilitating the grouping of metabolites into a processed data matrix across samples. Principal component analysis (PCA) was employed to identify potential batch effects, which were eliminated through linear mixed modelling.

VOCs were screened quantitatively by single-factor hypothesis test (SHT) and variable importance in the projection (VIP) via orthogonal partial least squares (OPLS) discriminant analysis (*p* < 0.05 & VIP >1). Differential VOCs were visualised using differential cluster plots. Random forest (RF), k-nearest neighbor (KNN), logistic regression (LR), XGBoost, and support vector machine (SVM), were employed to establish models. The sensitivity, specificity, and area under the curve (AUC) was calculated to evaluate model performance and receiver operating characteristic (ROC) curve were given for performance evaluation.

## 3. Results

### 3.1 Study population

Initially, 119 participants were recruited for LC and BC groups. Exclusion criteria were applied to participants aged outside the 18 to 80 years range, those who declined to participate, or those who provided invalid breath samples, resulting in a final cohort of 106 eligible participants for analysis. Of these, 85 were diagnosed with LC, and 21 were identified as BCs, as confirmed by histological assessments. Demographic and clinical data for these participants are presented in Table 1. Statistical comparisons between the case and control groups were performed on basic demographic characteristics, including age, gender, body mass index (BMI), smoking and alcohol consumption status, and the presence of underlying diseases. As detailed in Table 1, no significant differences were observed in these factors.

**Table 1.** Baseline data and statistical analysis of all enrolled participants.

| Groups |  | LC | BC | *P* values |
| --- | --- | --- | --- | --- |
| *n* |  | 85 | 21 |  |
| Age |  | 61.0 ± 9.9 | 54.4 ± 15.8 | 0.07 |
| BMI |  | 22.6 ± 4.1 | 23.6 ± 5.8 | 0.93 |
| Gender | Male | 31 (36.5%) | 10 (47.6%) | 0.35 |
|  | Female | 54 (63.5%) | 11 (52.4%) |  |
| Smoking | Never | 67 (78.8%) | 16 (76.2%) | 0.78 |
|  | Ever | 9 (10.6%) | 2 (9.5%) |  |
|  | Current | 9 (10.6%) | 3 (14.3%) |  |
| Alcohol | Never | 74 (87.1%) | 19 (90.5%) | 0.57 |
|  | Ever | 7 (8.2%) | 2 (9.5%) |  |
|  | Current | 4 (4.7%) | 0 (0.0%) |  |
| Underlying diseases | Yes | 13 (15.3%) | 5 (23.8%) | 0.35 |
|  | No | 72 (84.7%) | 16 (76.2%) |  |

Abbreviation: BC, benign controls; BMI, body mass index; LC, lung cancer.

### 3.2 VOC biomarkers

Following OPLS screening with criteria of *p* < 0.05 and VIP > 1, ten VOCs, including two unidentified entities, were consistently detected in the exhaled breath samples of both LC patients and BC groups. These compounds, alongside their discriminant values, are enumerated in Table 2. The majority of the identified VOCs were hydrocarbons, with the exception of hexamethylcyclotrisiloxane and acetophenone. Figure 1 presents a comparative analysis of the peak intensities of these VOCs between the two groups, highlighting a marked increase in LC patients, particularly for dodecane, methylcyclopentane, and the compound with a retention time of 3.692, which exhibited fold changes exceeding 10.

**Figure 1.**
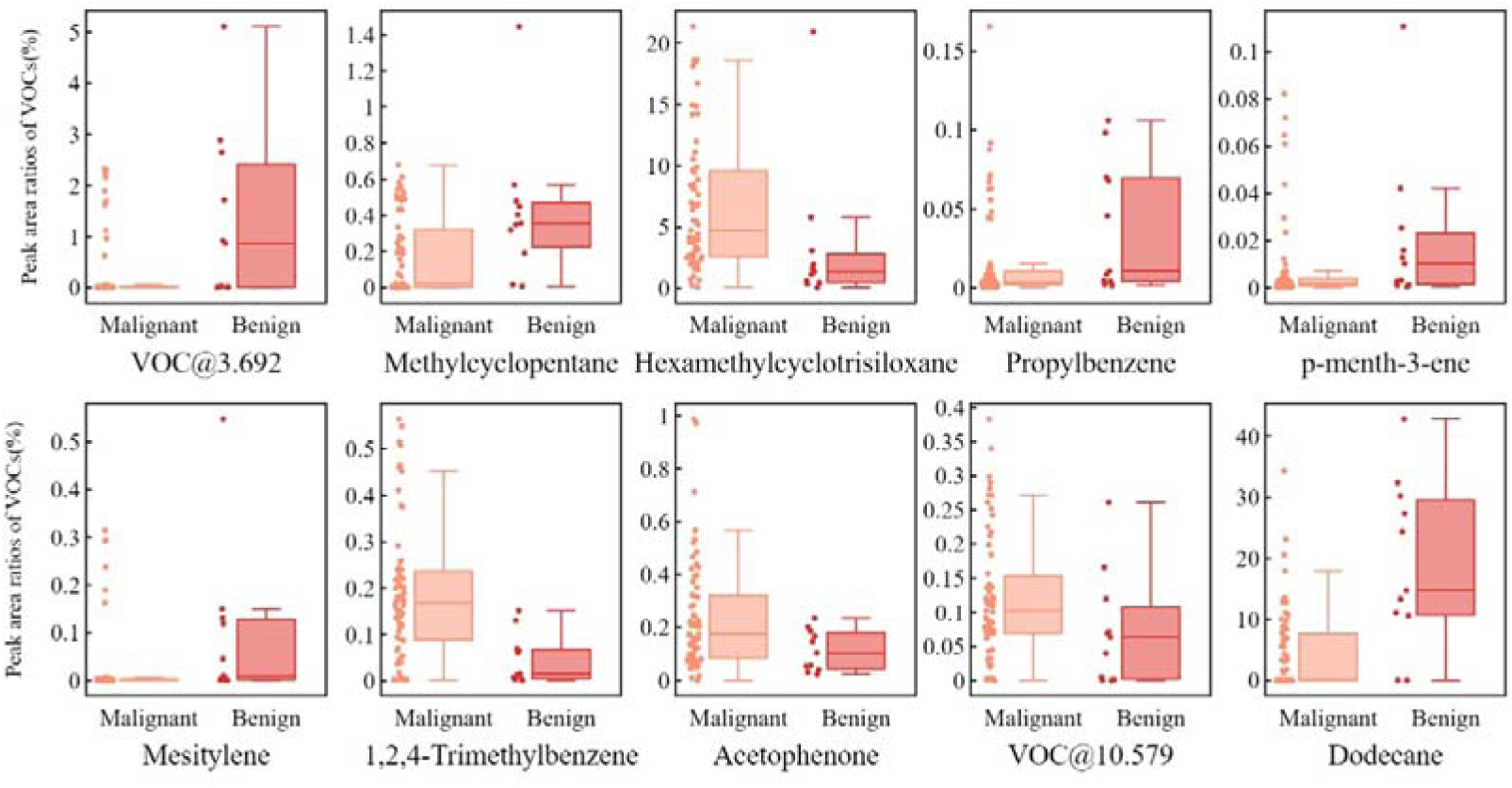
Box plots the differential VOC metabolites from μGC-μPID data.

**Table 2.**
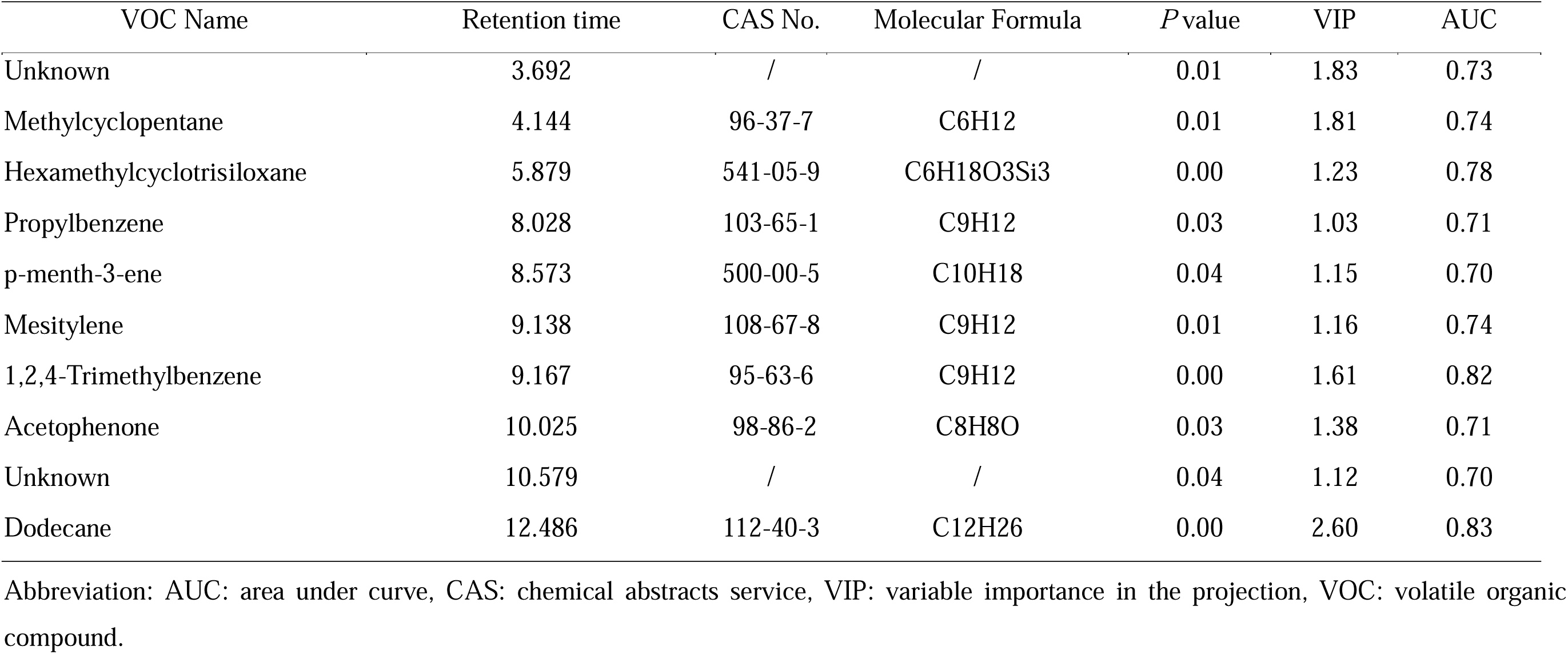
VOCs screened from μGC-μPID data.

| VOC Name | Retention time | CAS No. | Molecular Formula | *P* value | VIP | AUC |
| --- | --- | --- | --- | --- | --- | --- |
| Unknown | 3.692 | / | / | 0.01 | 1.83 | 0.73 |
| Methylcyclopentane | 4.144 | 96-37-7 | C<sub>6</sub>H<sub>12</sub> | 0.01 | 1.81 | 0.74 |
| Hexamethylcyclotrisiloxane | 5.879 | 541-05-9 | C<sub>6</sub>H<sub>18</sub>O<sub>3</sub>Si<sub>3</sub> | 0.00 | 1.23 | 0.78 |
| Propylbenzene | 8.028 | 103-65-1 | C<sub>9</sub>H<sub>12</sub> | 0.03 | 1.03 | 0.71 |
| p-menth-3-ene | 8.573 | 500-00-5 | C<sub>10</sub>H<sub>18</sub> | 0.04 | 1.15 | 0.70 |
| Mesitylene | 9.138 | 108-67-8 | C<sub>9</sub>H<sub>12</sub> | 0.01 | 1.16 | 0.74 |
| 1,2,4-Trimethylbenzene | 9.167 | 95-63-6 | C<sub>9</sub>H<sub>12</sub> | 0.00 | 1.61 | 0.82 |
| Acetophenone | 10.025 | 98-86-2 | C<sub>8</sub>H<sub>8</sub>O | 0.03 | 1.38 | 0.71 |
| Unknown | 10.579 | / | / | 0.04 | 1.12 | 0.70 |
| Dodecane | 12.486 | 112-40-3 | C<sub>12</sub>H<sub>26</sub> | 0.00 | 2.60 | 0.83 |

Abbreviation: AUC: area under curve, CAS: chemical abstracts service, VIP: variable importance in the projection, VOC: volatile organic compound.

### 3.3 Model performance

The individual diagnostic efficacy of the aforementioned differential VOCs for LC is delineated in Table 2. Diagnostic models (Model Group 1) incorporating these ten VOCs from the OPLS screening were evaluated using five distinct machine learning (ML) algorithms. These models achieved AUCs ranging from 0.79 to 0.90, sensitivities from 0.95 to 1.00, specificities from 0.84 to 0.88, and accuracies from 0.84 to 0.86, as detailed in Table 3, along with F1 scores and accuracy indices. Furthermore, to enhance or maintain the predictive performance and robustness of the model, a smaller subset of seven VOCs was incrementally refined through recursive feature elimination with cross-validation (RFECV) and RF feature-selection techniques, excluding methylcyclopentane, acetophenone, and the aforementioned unknown compound VOC@10.579. Diagnostic models (Model Group 2) incorporating the final seven VOCs are showcased in Table 4. Notably, the KNN algorithm demonstrated the most proficient performance (AUC > 0.9) among the ML models in both groups, with its ROC curve depicted in Figure 2 in Model Group 2.

**Figure 2.**
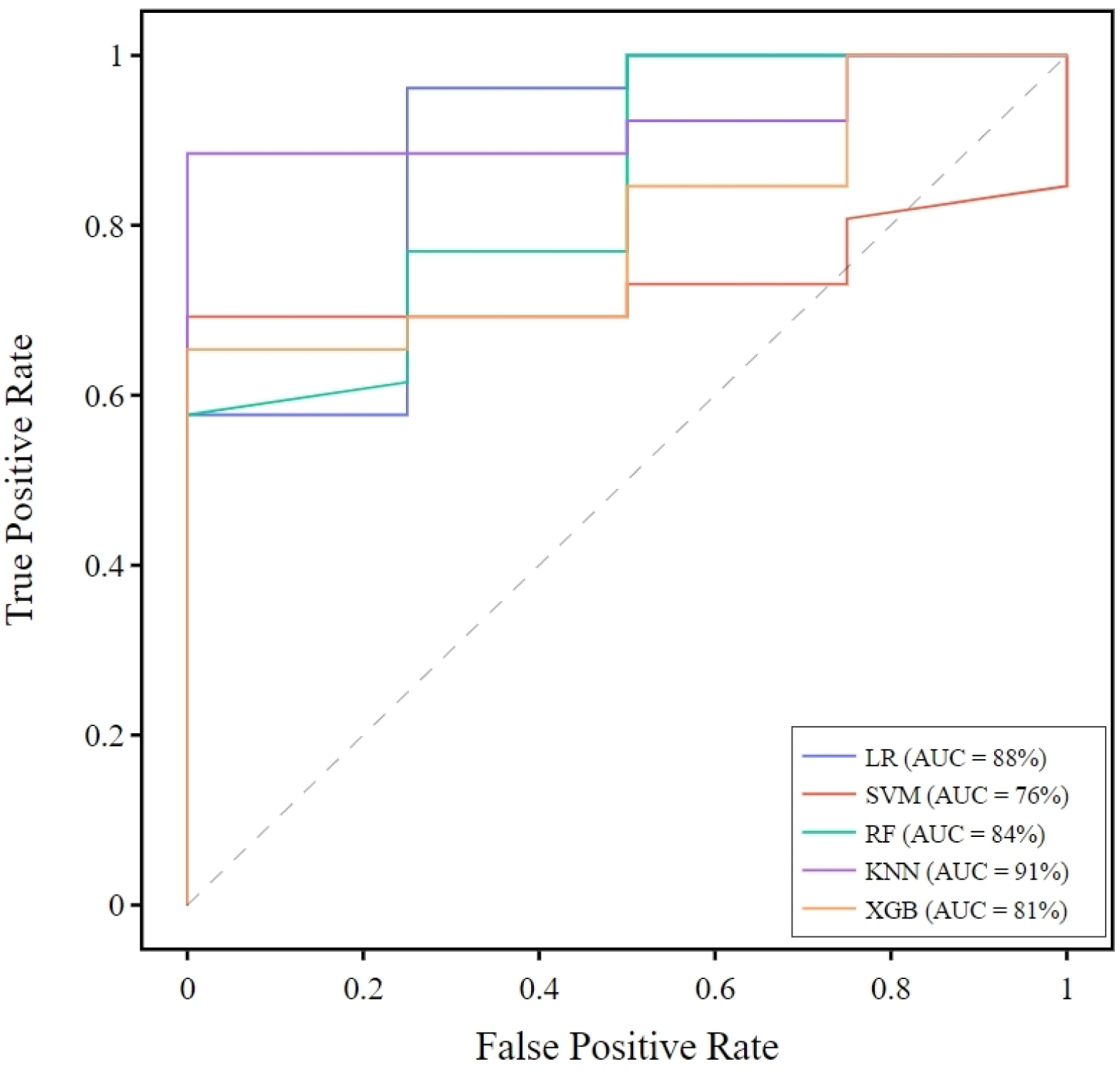
ROC curve of the KNN model in Model group 2. AUC: area under curve, KNN: k-nearest neighbor, LR: logistic regression, RF: random forest, SVM: support vector machine, XGB: XGBoost.

**Table 3.** Model performances from μGC-μPID data with 10 statistically different VOCs (Model group 1) and 7 model-selected VOCs (Model group 2).

| Algorithms | Model group 1 |  |  |  |  | Model group 2 |  |  |  |  |
| --- | --- | --- | --- | --- | --- | --- | --- | --- | --- | --- |
|  | AUC | F1 | Accuracy | Sensitivity | Specificity | AUC | F1 | Accuracy | Sensitivity | Specificity |
| LR | 0.86±0.02 | 0.92±0.01 | 0.86±0.02 | 0.97±0.01 | 0.88±0.02 | 0.87±0.02 | 0.92±0.01 | 0.87±0.02 | 0.97±0.01 | 0.88±0.02 |
| SVM | 0.77±0.06 | 0.91±0.01 | 0.84±0.02 | 1.00±0.00 | 0.84±0.02 | 0.77±0.07 | 0.91±0.01 | 0.84±0.02 | 0.96±0.02 | 0.87±0.02 |
| RF | 0.85±0.04 | 0.92±0.01 | 0.85±0.02 | 0.98±0.01 | 0.86±0.02 | 0.84±0.04 | 0.91±0.02 | 0.85±0.03 | 0.97±0.02 | 0.86±0.02 |
| KNN | 0.90±0.02 | 0.91±0.01 | 0.84±0.02 | 1.00±0.00 | 0.84±0.02 | 0.91±0.02 | 0.91±0.01 | 0.84±0.02 | 1.00±0.00 | 0.84±0.02 |
| XGBoost | 0.79±0.05 | 0.90±0.02 | 0.83±0.03 | 0.95±0.02 | 0.87±0.03 | 0.80±0.04 | 0.90±0.02 | 0.83±0.03 | 0.95±0.02 | 0.87±0.02 |

Abbreviation: AUC, area under curve; KNN, k-nearest neighbor; LR, logistic regression; RF, random forest; SVM, support vector machine.

**Table 4.** Accuracy of 7-VOC models on the classification of patients with recurrence, metastasis and cancer sites.

| Patients | LR | SVM | RF | XGBoost | KNN |
| --- | --- | --- | --- | --- | --- |
| NPTML #1 | 1 | 1 | 1 | 1 | 1 |
| NPTML #2 | 1 | 1 | 1 | 1 | 1 |
| LCPS | 1 | 0 | 0 | 0 | 1 |
| OC #1 | 0 | 0 | 0 | 0 | 1 |
| OC #2 | 0 | 0 | 0 | 1 | 1 |
| OC #3 | 1 | 0 | 0 | 1 | 1 |

Abbreviation: KNN, k-nearest neighbor; LCPS, lung cancer post-surgery; LR, logistic regression; NPTML, non-pulmonary tumours metastasised to the lung; OC, oesophageal cancer; RF, random forest; SVM, support vector machine.

### 3.4 Influence of recurrence, metastasis and cancer sites

Three additional patient groups were also evaluated with the aforementioned 7-VOC model: (1) patients with recurrent LC post-surgery, (2) patients with non-pulmonary tumours metastasised to the lung, and (3) patients with oesophageal cancer. The results, as presented in Table 5, indicated that the KNN model achieved 100% accuracy in classifying all groups into the LC category.

## 4. Discussion

This proof-of-concept study represents the inaugural investigation into the utility of μGC-μPID for differentiating LC patients from BCs among CT abnormal subjects. The models, trained and evaluated in a blinded fashion, achieved sensitivities of 0.95 to 1.00, specificities of 0.84 to 0.88, and AUCs of 0.77 to 0.91, utilising seven potential VOC biomarkers, six of which were identified. Furthermore, analysis of the influence of recurrence, metastasis, and cancer sites suggested that these VOCs are not LC-specific but rather indicative of malignant tumours with a high degree of probability. These findings underscore the potential diagnostic value of breath VOCs in LC and lay the groundwork for the clinical application of μGC-μPID breath analysis technology.

Among the six identified VOC biomarkers, all have been previously linked to various cancers. For instance, hexamethylcyclotrisiloxane, a common environment pollutant, which was supposed to originate from background emissions of the thermal desorption process or emollients [43], affects expressions of BRCA1, BRCA2, CHEK1 and CHEK1 mRNA [44]. It was found to exist in SW620 CRC cells with a different average level compared to those in normal cells [45]. Dodecane is a carcinogen mainly absorbed by inhalation and metabolized by the liver microsomal mixed-function oxidase system. It has been consistently reported as a breath VOC marker for LC [46–49], CRC [50] and gastric cancer [51]. Wang et al. also identified it as a characteristic VOC in LC tissue compared to adenocarcinoma, squamous carcinoma, and SCLC cell lines [19]. Propylbenzene has been found to increase in the exhaled air of LC patients relative to healthy controls [49, 52], whereas our study observed a decreased concentration. It is located in human cell membranes and involved in oxidation and hydroxylation pathways. 1,2,4-trimethylbenzene has been reported as a breath biomarker for LC by Phillips et al. [29] and Chen et al. [52], and is also implicated in the urinary discrimination of oncological groups, including leukaemia, colorectal, and lymphoma, from healthy individuals [53]. Mesitylene, a natural product found in Carica papaya, has been noted as a characteristic skin VOC in a case report of malignant melanoma [54] and was reported to excreted unchanged by the lungs. P-menth-3-ene is associated with oxidative stress, a common feature in neoplastic diseases [55]. It is also found in Angelica gigas, a medicinal herb showing potential anti-cancer effects [56].

Our study diverges from existing work employing GC-MS, online MS, or eNose instruments, not only due to the distinct combination of VOC biomarkers identified but also the methodology. The μGC-μPID utilised in this study is a rapid, point-of-care (POC), non-target breath VOC analyser capable of direct breath sampling with exceptionally low detection limits, typically 10 parts per trillion (ppt). This technology bypasses the time-consuming and complex procedures of traditional GC-MS, the limited receptive range and lack of qualitative or quantitative capabilities of eNose, thereby facilitating a more practical and adaptable clinical translation.

Nevertheless, several limitations are acknowledged. Most notably, the study’s sample size is modest, reflecting a single-centre pilot study. Further research and validation are essential to refine a more consistent and precise panel of VOCs with a larger and multi-centric cohort. Secondly, an evaluation of LC stages was not conducted due to the limited number of participants, a shortcoming that is currently being addressed. Thirdly, the metabolic pathways of the potential biomarkers remain poorly understood, which diminishes the clinical persuasiveness of breath VOC diagnosis without clear etiologies and metabolic mechanisms. Further foundational biological and medical research is imperative for the field of breathomics. Lastly, the μGC-μPID breath analyser’s capacity to detect a limited range of VOCs, in comparison to traditional GC-MS, is constrained by the selected internal materials, length, and operating temperatures of the columns, as well as the 10.6 eV ionisation potential. Ongoing development of the hardware aims to enhance separation and detection capabilities.

In conclusion, this study presents the development and evaluation of a rapid POC breath test for the differential diagnosis of LC from benign lung diseases in CT abnormal groups, employing our self-developed μGC-μPID breath analyser. The results indicate that the proposed breath VOC biomarkers and methods can accurately discriminate LC from control groups, with the model extending favourably to the recurrence and metastasis of pulmonary cancer and oesophageal cancer. Six potential VOC biomarkers were identified for LC differential diagnosis. Further analysis on subdivided group differentiation and extensive cohort studies are warranted prior to clinical application.

## Data Availability

Data sharing is not applicable to this article as no datasets were generated or analyzed during the current study.

## References

1. Sung, H., et al., Global Cancer Statistics 2020: GLOBOCAN Estimates of Incidence and Mortality Worldwide for 36 Cancers in 185 Countries. CA Cancer J Clin, 2021. 71(3): p. 209–249.

2. Thandra, K.C., et al., Epidemiology of lung cancer. Contemp Oncol (Pozn), 2021. 25(1): p. 45–52.

3. Li, G., et al., Metal Oxide Semiconductor Gas Sensors for Lung Cancer Diagnosis. Chemosensors, 2023. 11(4): p. 251.

4. Guerrera, F., et al., Exploring Stage I non-small-cell lung cancer: development of a prognostic model predicting 5-year survival after surgical resection†. Eur J Cardiothorac Surg, 2015. 47(6): p. 1037–43.

5. Ibrahim, W., et al., Breathomics for the clinician: the use of volatile organic compounds in respiratory diseases. Thorax, 2021. 76(5): p. 514–521.

6. Amann, A., et al., Analysis of Exhaled Breath for Disease Detection. Annual Review of Analytical Chemistry, 2014. 7(1): p. 455–482.

7. Jia, Z., et al., Breath Analysis for Lung Cancer Early Detection—A Clinical Study. Metabolites, 2023. 13(12): p. 1197.

8. Koureas, M., et al., Comparison of Targeted and Untargeted Approaches in Breath Analysis for the Discrimination of Lung Cancer from Benign Pulmonary Diseases and Healthy Persons. Molecules, 2021. 26(9): p. 2609.

9. Liu, B., et al., Lung cancer detection via breath by electronic nose enhanced with a sparse group feature selection approach. Sensors and Actuators B: Chemical, 2021. 339: p. 129896.

10. Binson, V.A., et al., Prediction of Pulmonary Diseases With Electronic Nose Using SVM and XGBoost. IEEE Sensors Journal, 2021. 21(18): p. 20886–20895.

11. Koureas, M., et al., Target Analysis of Volatile Organic Compounds in Exhaled Breath for Lung Cancer Discrimination from Other Pulmonary Diseases and Healthy Persons. Metabolites, 2020. 10(8): p. 317.

12. Wang, M., et al., Confounding effect of benign pulmonary diseases in selecting volatile organic compounds as markers of lung cancer. Journal of Breath Research, 2018. 12(4): p. 046013.

13. Binson, V.A., M. Subramoniam, and L. Mathew, Discrimination of COPD and lung cancer from controls through breath analysis using a self-developed e-nose. Journal of Breath Research, 2021. 15(4): p. 046003.

14. Tirzīte, M., et al., Detection of lung cancer in exhaled breath with an electronic nose using support vector machine analysis. Journal of Breath Research, 2017. 11(3): p. 036009.

15. Schumer, E.M., et al., High sensitivity for lung cancer detection using analysis of exhaled carbonyl compounds. J Thorac Cardiovasc Surg, 2015. 150(6): p. 1517–22; discussion 1522-4.

16. Bousamra, M., 2nd, et al., Quantitative analysis of exhaled carbonyl compounds distinguishes benign from malignant pulmonary disease. J Thorac Cardiovasc Surg, 2014. 148(3): p. 1074–80; discussion 1080-1.

17. Liu, H., et al., Characterization of Volatile Organic Metabolites in Lung Cancer Pleural Effusions by SPME–GC/MS Combined with an Untargeted Metabolomic Method. Chromatographia, 2014. 77(19): p. 1379–1386.

18. Broza, Y.Y., et al., A nanomaterial-based breath test for short-term follow-up after lung tumor resection. Nanomedicine: Nanotechnology, Biology and Medicine, 2013. 9(1): p. 15–21.

19. Wang, Y., et al., The analysis of volatile organic compounds biomarkers for lung cancer in exhaled breath, tissues and cell lines. Cancer Biomark, 2012. 11(4): p. 129–37.

20. Ding, X., et al., Diagnosis of primary lung cancer and benign pulmonary nodules: a comparison of the breath test and 18F-FDG PET-CT. Frontiers in Oncology, 2023. 13.

21. Rai, S.N., et al., Multigroup prediction in lung cancer patients and comparative controls using signature of volatile organic compounds in breath samples. PLOS ONE, 2022. 17(11): p. e0277431.

22. Chen, X., et al., Calculated indices of volatile organic compounds (VOCs) in exhalation for lung cancer screening and early detection. Lung Cancer, 2021. 154: p. 197–205.

23. Phillips, M., T.L. Bauer, and H.I. Pass, A volatile biomarker in breath predicts lung cancer and pulmonary nodules. J Breath Res, 2019. 13(3): p. 036013.

24. Shlomi, D., et al., Detection of Lung Cancer and EGFR Mutation by Electronic Nose System. J Thorac Oncol, 2017. 12(10): p. 1544–1551.

25. Li, M., et al., Breath carbonyl compounds as biomarkers of lung cancer. Lung Cancer, 2015. 90(1): p. 92–7.

26. Fu, X.A., et al., Noninvasive detection of lung cancer using exhaled breath. Cancer Med, 2014. 3(1): p. 174–81.

27. Peled, N., et al., Non-invasive breath analysis of pulmonary nodules. J Thorac Oncol, 2012. 7(10): p. 1528–33.

28. Gordon, S.M., et al., Volatile organic compounds in exhaled air from patients with lung cancer. Clin Chem, 1985. 31(8): p. 1278–82.

29. Phillips, M., et al., Volatile organic compounds in breath as markers of lung cancer: a cross-sectional study. The Lancet, 1999. 353(9168): p. 1930–1933.

30. Phillips, M., et al., Detection of lung cancer using weighted digital analysis of breath biomarkers. Clin Chim Acta, 2008. 393(2): p. 76–84.

31. Phillips, M., et al., Blinded Validation of Breath Biomarkers of Lung Cancer, a Potential Ancillary to Chest CT Screening. PLOS ONE, 2015. 10(12): p. e0142484.

32. Ulanowska, A., et al., The application of statistical methods using VOCs to identify patients with lung cancer. J Breath Res, 2011. 5(4): p. 046008.

33. Ligor, T., Ł. Pater, and B. Buszewski, Application of an artificial neural network model for selection of potential lung cancer biomarkers. Journal of Breath Research, 2015. 9(2): p. 027106.

34. Corradi, M., et al., Exhaled breath analysis in suspected cases of non-small-cell lung cancer: a cross-sectional study. Journal of Breath Research, 2015. 9(2): p. 027101.

35. Schmidt, F., et al., Mapping the landscape of lung cancer breath analysis: A scoping review (ELCABA). Lung Cancer, 2023. 175: p. 131–140.

36. Hanna, G.B., et al., Accuracy and Methodologic Challenges of Volatile Organic Compound-Based Exhaled Breath Tests for Cancer Diagnosis: A Systematic Review and Meta-analysis. JAMA Oncol, 2019. 5(1): p. e182815.

37. Wang, J., et al., Belt-Mounted Micro-Gas-Chromatograph Prototype for Determining Personal Exposures to Volatile-Organic-Compound Mixture Components. Anal Chem, 2019. 91(7): p. 4747–4754.

38. Sharma, R., et al., Portable Breath-Based Volatile Organic Compound Monitoring for the Detection of COVID-19 During the Circulation of the SARS-CoV-2 Delta Variant and the Transition to the SARS-CoV-2 Omicron Variant. JAMA Netw Open, 2023. 6(2): p. e230982.

39. Sharma, R., et al., Real Time Breath Analysis Using Portable Gas Chromatography for Adult Asthma Phenotypes. Metabolites, 2021. 11(5): p. 265.

40. Zhou, M., et al., Rapid breath analysis for acute respiratory distress syndrome diagnostics using a portable two-dimensional gas chromatography device. Analytical and Bioanalytical Chemistry, 2019. 411(24): p. 6435–6447.

41. Ruchi, S., et al., Breath analysis for detection and trajectory monitoring of acute respiratory distress syndrome in swine. ERJ Open Research, 2022. 8(1): p. 00154–2021.

42. Picciariello, A., et al., Colorectal Cancer Diagnosis through Breath Test Using a Portable Breath Analyzer—Preliminary Data. Sensors, 2024. 24(7): p. 2343.

43. Zuo, Z., et al., VOC Outgassing from Baked and Unbaked Ventilation Filters. Aerosol and Air Quality Research, 2010. 10(3): p. 265–271.

44. Farasani, A. and P.D. Darbre, Exposure to cyclic volatile methylsiloxanes (cVMS) causes anchorage-independent growth and reduction of BRCA1 in non-transformed human breast epithelial cells. J Appl Toxicol, 2017. 37(4): p. 454–461.

45. Wang, G., et al., Determination of volatile organic compounds in SW620 colorectal cancer cells and tumor-bearing mice. Journal of Pharmaceutical and Biomedical Analysis, 2019. 167: p. 30–37.

46. Peng, G., et al., Detection of lung, breast, colorectal, and prostate cancers from exhaled breath using a single array of nanosensors. British Journal of Cancer, 2010. 103(4): p. 542–551.

47. Filipiak, W., et al., Comparative analyses of volatile organic compounds (VOCs) from patients, tumors and transformed cell lines for the validation of lung cancer-derived breath markers. Journal of Breath Research, 2014. 8(2): p. 027111.

48. Handa, H., et al., Exhaled Breath Analysis for Lung Cancer Detection Using Ion Mobility Spectrometry. PLOS ONE, 2014. 9(12): p. e114555.

49. Schallschmidt, K., et al., Comparison of volatile organic compounds from lung cancer patients and healthy controls—challenges and limitations of an observational study. Journal of Breath Research, 2016. 10(4): p. 046007.

50. Wang, C., et al., Noninvasive detection of colorectal cancer by analysis of exhaled breath. Analytical and bioanalytical chemistry, 2014. 406: p. 4757–4763.

51. Chen, Y., et al., Breath analysis based on surface-enhanced Raman scattering sensors distinguishes early and advanced gastric cancer patients from healthy persons. ACS nano, 2016. 10(9): p. 8169–8179.

52. Xing, C., et al. A Non-invasive Detection of Lung Cancer Combined Virtual Gas Sensors Array with Imaging Recognition Technique. in 2005 IEEE Engineering in Medicine and Biology 27th Annual Conference. 2005.

53. Silva, C.L., M. Passos, and J.S. Câmara, Investigation of urinary volatile organic metabolites as potential cancer biomarkers by solid-phase microextraction in combination with gas chromatography-mass spectrometry. British Journal of Cancer, 2011. 105(12): p. 1894–1904.

54. Abaffy, T., et al., A case report-Volatile metabolomic signature of malignant melanoma using matching skin as a control. Journal of cancer science & therapy, 2011. 3(6): p. 140.

55. Toyokuni, S., Molecular mechanisms of oxidative stressLJinduced carcinogenesis: From epidemiology to oxygenomics. IUBMB life, 2008. 60(7): p. 441–447.

56. Kim, S.H., et al., Anti-cancer activity of Angelica gigas by increasing immune response and stimulating natural killer and natural killer T cells. BMC Complement Altern Med, 2018. 18(1): p. 218.

